# Development and validation of multivariable machine learning algorithms to predict risk of cancer in symptomatic patients referred urgently from primary care

**DOI:** 10.1101/2020.10.23.20218198

**Authors:** Richard S Savage, Mike Messenger, Richard D Neal, Rosie Ferguson, Colin Johnston, Katherine L Lloyd, Matthew D Neal, Nigel Sansom, Peter Selby, Nisha Sharma, Bethany Shinkins, Jim R Skinner, Giles Tully, Sean Duffy, Geoff Hall

**Author notes:** Joint lead author. Joint last author. Corresponding author (Richard S Savage,).

## Abstract

**Background:** Urgent Suspected Cancer (Two Week Wait, 2WW) referrals have improved early cancer detection but are increasingly a major burden on NHS services. This has been exacerbated by the COVID-19 pandemic.

**Method:** We developed and validated tests to assess the risk of any cancer for 2WW patients. The tests use routine blood measurements (FBC, U&E, LFTs, tumour markers), combining them using machine learning and statistical modelling. Algorithms were developed and validated for nine 2WW pathways using retrospective data from 371,799 referrals to Leeds Teaching Hospitals Trust (development set 224,669 referrals, validation set 147,130 referrals). A minimum set of blood measurements were required for inclusion, and missing data were modelled internally by the algorithms.

**Results:** We present results for two clinical use-cases. In use-case 1, the algorithms identify 20% of patients who do not have cancer and may not need an urgent 2WW referral. In use-case 2, they identify 90% of cancer cases with a high probability of cancer that could be prioritised for review.

**Conclusions:** Combining a panel of widely available blood markers produces effective blood tests for cancer for NHS 2WW patients. The tests are affordable, can be deployed rapidly to any NHS pathology laboratory with no additional hardware requirements.

## 1 Background

A major NHS cancer policy to diagnose cancer earlier led to the introduction of Urgent Suspected Cancer referrals. These referrals are predicated on the risk of symptomatic patients having cancer.^1^ Trusts assess patients within two weeks (‘two-week wait’ (2WW) referral). The 2WW pathways have contributed to improving outcomes; higher general practice use of referrals for suspected cancer is associated with lower mortality for the four most common types of cancer (prostate, breast, lung, and colorectal).^2^

This approach places a major strain on diagnostic services on NHS England, with over 2 million 2WW referrals annually, and a 10% year-on-year increase in referrals over the past decade.^3^ This highlights an unsustainable burden on existing services, workforce and financial resources. Whilst there is variation between cancer pathways, only 7% overall of 2WW referral patients are diagnosed with cancer.^3^ Many patients are therefore subject to unnecessary psychological distress, as well as being exposed to diagnostic tests which may inadvertently cause harm. Clearly there is a need to improve the efficiency of these pathways.

These challenges are exacerbated by the current COVID-19 crisis. The NHS capacity to assess 2WW referrals is reduced, and a backlog of referrals continues to build.^3,4^ These unprecedented challenges urgently require new solutions. COVID-19 has presented an opportunity for GPs to permanently change how they use emerging technologies.^5^

Many biomarkers have been evaluated for their use in cancer diagnosis; however only a few are currently used in either primary or secondary care settings. A systematic mapping review identified 94 ctDNA studies alone, highlighting how much more work is required prior to clinical use.^6^ Companies like GRAIL and Freenome are pursuing this, with clinical trials ongoing.^7,8^ There is also evidence that signals from a range of different analytes can be usefully combined via machine learning.^9^

Using such approaches to triage cancer referrals should bring benefits to patients, health-systems and the economy. For example, a *rule-out* test for symptomatic patients, like those referred to the NHS 2WW, could identify those with very low cancer risk, allowing many patients without cancer to avoid unnecessary procedures and freeing up diagnostic capacity for those at greater risk.

The work presented in this paper addresses the top three priority areas identified by Badrick et al (2019), including: a simple, non-invasive, painless and convenient test to detect cancer early; a blood test to detect some or all cancers early that can be included into routine care; and a test that is easily accessible to General Practice.^10^

We report the development and validation of a set of machine learning algorithms to provide a calibrated risk probability of cancer (a score between zero and one, higher values indicating greater risk of cancer) for triaging symptomatic patients. A calibrated risk probability has a variety of clinical uses. This paper focuses on the two use-cases for the NHS 2WW:

Use-Case 1 - a rule-out test when patient has a very low risk of cancer, allowing initial management in primary care.

Use-Case 2 - a way of identifying patients at high risk of having cancer to fast-track them for further tests.

## 2 Methods

### Methodological Design and Source of Data

This work is a single centre, retrospective diagnostic prediction study (classified as a Type 2b study by the TRIPOD statement.^11^ The prediction algorithms were developed and validated on a large data set from a single geographic area, split chronologically into two independent cohorts.

The data set contained 371,799 consecutive 2WW referrals in the Leeds region from 2011-2019. The development cohort was composed of 224,669 consecutive patients with an urgent suspected cancer referral in Leeds between January 2011 and December 2016. The diagnostic algorithms developed were then externally validated on a similar consecutive sample of 147,130 patients (between January 2017 and December 2019). Both development and validation sets were selected using the same inclusion and exclusion criteria and both received the same pre-processing, consisting of removing greater-than (“>“) symbols from blood analyte values in the data, and setting data values with less-than (“<“) values to zero. This is a simple imputation for the case where a pathology laboratory returns a result outside the reportable range. Because the chosen machine learning algorithms are not sensitive to scaling of individual variables, it was not necessary to normalise the inputs.

### 2.1 Participants

Patients were selected because they received a 2WW referral to Leeds Teaching Hospitals NHS Trust during the above timeframe. Referrals were included for all 2WW pathways, and all patients over the age of 18 with a minimum set of blood counts and biochemistry measurements available were included in the cohort. Occasional multiple referrals of the same patient (for example to different 2WW pathways) is expected in this data set – such instances are infrequent. Patients from all 2WW pathways were included in the development set; patients from the nine 2WW pathways at LTHT considered in this paper were included in the validation set. Validation was restricted to these nine 2WW pathways (which account for ∼98% of all 2WW referrals in England) because the remaining pathways, being much smaller, did not have sufficient validation data to provide useful validation. Patients not fulfilling these criteria were excluded from the analysis. All patients were followed up to 12 months after the conclusion of their referral, or until February 2020. Patients in the validation set (i.e. referred from January 2017 onwards) only required the outcome of the 2WW referral and therefore the possibility of censoring of outcomes up to 12 months did not affect the validation results.

### 2.2 Outcome

The algorithms were trained to predict whether or not a patient would receive a cancer diagnosis. Outcome labels were derived from ICD10 diagnostic codes from the Leeds secondary care cancer clinical database. ‘Cancer’ was defined as any patient diagnosed with a malignant (ICD10 ‘C’ codes) or in situ (appropriate subset of ICD10 ‘D’ codes) neoplasm as the result of their referral or within the subsequent 12-month period for the purposes of model development. Diagnoses as the result of an urgent referral were used as outcomes in the validation analyses, to match the intended clinical setting. Benign neoplasms were defined as ‘Not Cancer’. The full list of ICD10 codes designated as ‘cancer’ are in the supplementary materials.

### 2.3 Predictors

The variables for each patient include a full blood count, a range of biochemistry measurements, a panel of standard tumour markers, plus age and sex. All predictors were included on their natural scale (i.e. they were not normalised or dichotomised).

As a retrospective cohort, blood measurements were used where they were available in the database up to 90 days prior to referral or up to 14 days post referral. This was done to seek a reasonable balance between missing data and possible bias (for example if blood measurements were made after a diagnosis had been established). For example, it is risky to use blood measurements taken more than 14 days post-referral as there is an increasing chance that those bloods could have been ordered by a clinician in response to a confirmed diagnosis of cancer. In routine clinical use, all model predictors would be available at the time.

### 2.4 Sample Size

The protocol stated the design as predicated on a goal of achieving a Negative Predictive Value (NPV) of 0.99 or greater. If we assume that we would like to determine the size of the distance from the 2.5% centile of the NPV to the point estimate (i.e. the distance between the lower bound of the 95% confidence interval (CI) and the point estimate), we can therefore determine the number of patients required in the denominator of the NPV calculation. For a 0.05 lower CI size, we require 100 patients in the denominator; for a 0.02 lower CI size we require 300 patients in the denominator. With a design goal of achieving 20% rule-out rate, this would therefore require approximately (100)/(0.2) = 500 total cases per pathway for a 0.05 lower CI size, or (300)/(0.2) = 1500 total cases per pathway for a 0.02 lower CI size.

### 2.5 Management of Missing Data

Missing data is a key issue for this cohort as many patients did not have bloods in this timeframe (see Tables 1, 2). Patients were identified who had full blood counts and a minimum subset of biochemistry data, and this subset was used to train the algorithms. The core algorithms use a gradient boosting model including an inbuilt method for imputing missing data which infers from the data how to handle missing data values, by learning at each decision tree node in the ensemble which branch a missing value should be assigned to. Early work during model development showed that this inbuilt method modestly outperformed (in a statistical sense) simple imputation methods, and has the advantage of simplifying the model development somewhat.

**Table 1:**
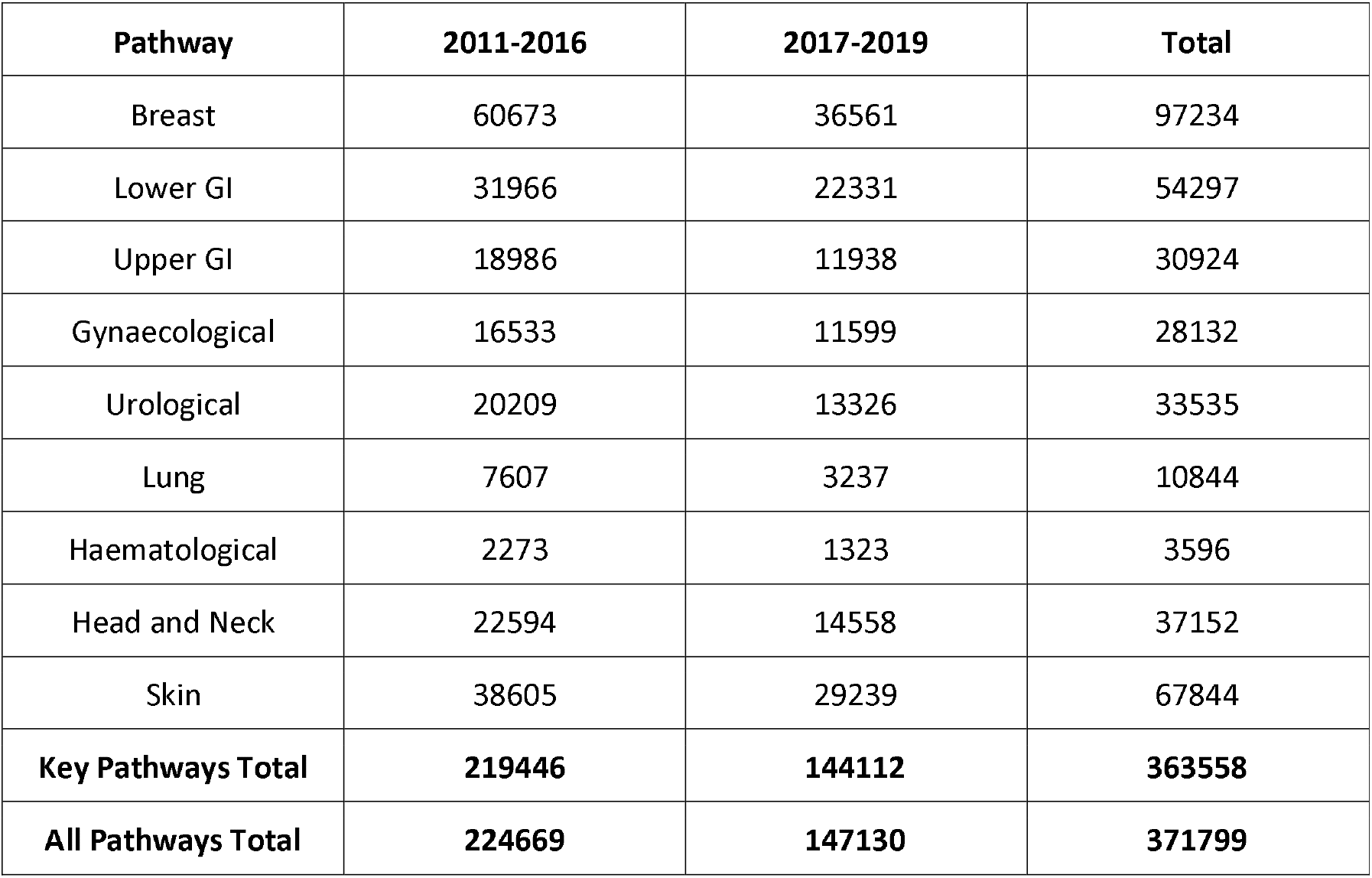
Total Number of Cases per Pathway (2011-2019)

| Pathway | 2011-2016 | 2017-2019 | Total |
| --- | --- | --- | --- |
| Breast | 60673 | 36561 | 97234 |
| Lower GI | 31966 | 22331 | 54297 |
| Upper GI | 18986 | 11938 | 30924 |
| Gynaecological | 16533 | 11599 | 28132 |
| Urological | 20209 | 13326 | 33535 |
| Lung | 7607 | 3237 | 10844 |
| Haematological | 2273 | 1323 | 3596 |
| Head and Neck | 22594 | 14558 | 37152 |
| Skin | 38605 | 29239 | 67844 |
| **Key Pathways Total** | **219446** | **144112** | **363558** |
| **All Pathways Total** | **224669** | **147130** | **371799** |

**Table 2:**
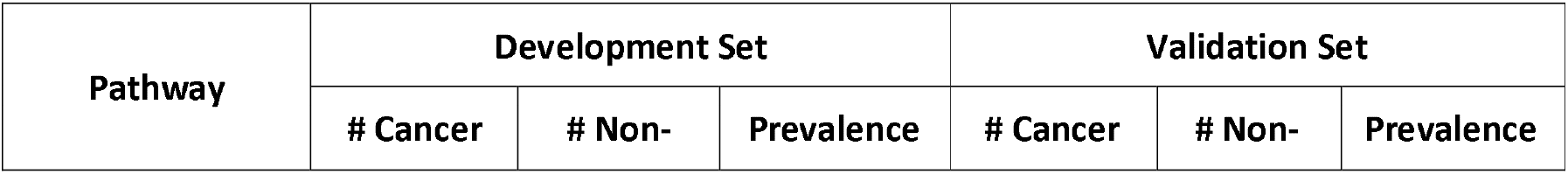
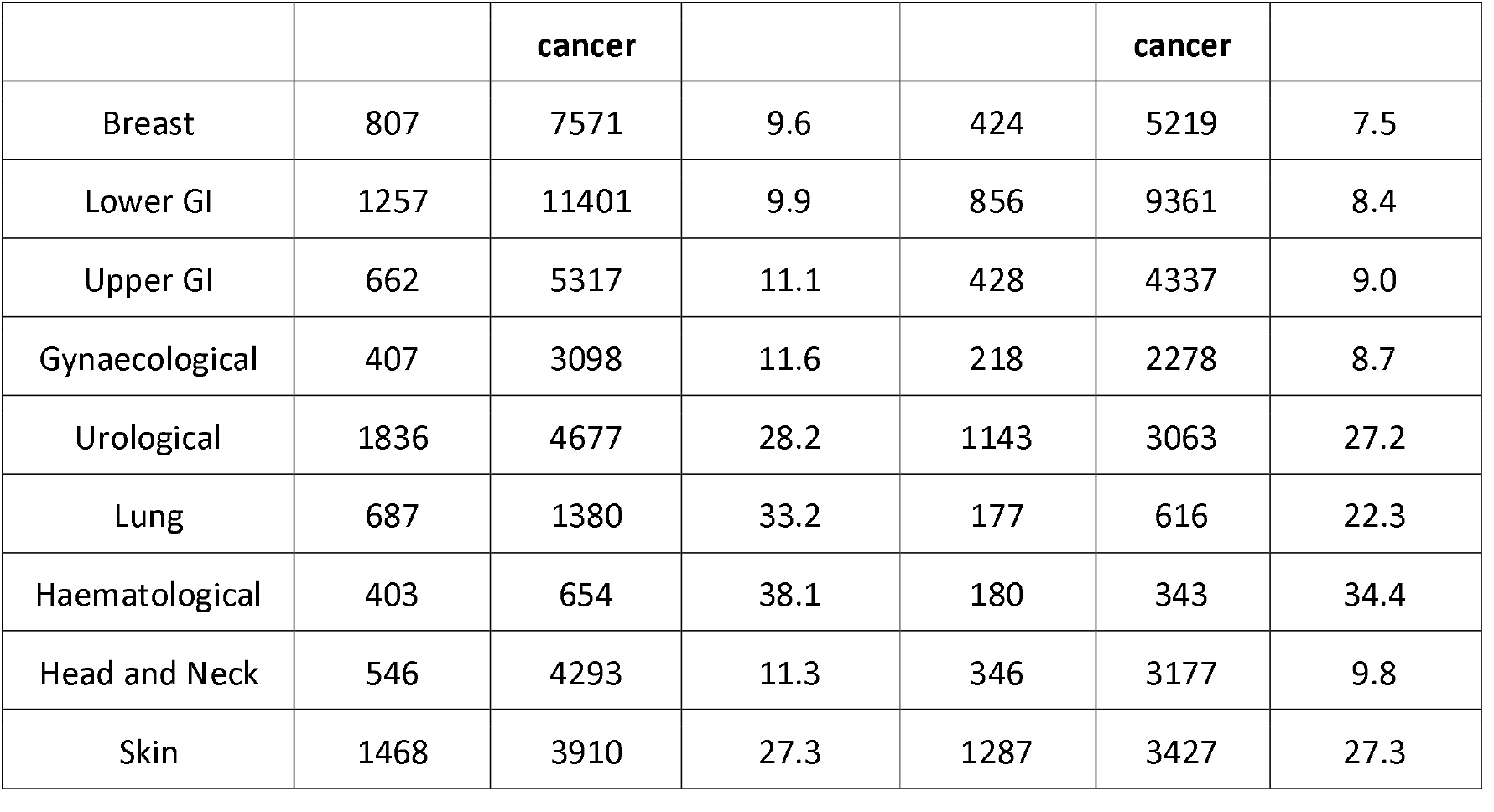
Number of Cases Meeting Bloods Criteria

| Pathway | Development Set |  |  | Validation Set |  |  |
| --- | --- | --- | --- | --- | --- | --- |
|  | # Cancer | # Non- | Prevalence | # Cancer | # Non- | Prevalence |

|  |  | **cancer** |  |  | **cancer** |  |
| --- | --- | --- | --- | --- | --- | --- |
| Breast | 807 | 7571 | 9.6 | 424 | 5219 | 7.5 |
| Lower GI | 1257 | 11401 | 9.9 | 856 | 9361 | 8.4 |
| Upper GI | 662 | 5317 | 11.1 | 428 | 4337 | 9.0 |
| Gynaecological | 407 | 3098 | 11.6 | 218 | 2278 | 8.7 |
| Urological | 1836 | 4677 | 28.2 | 1143 | 3063 | 27.2 |
| Lung | 687 | 1380 | 33.2 | 177 | 616 | 22.3 |
| Haematological | 403 | 654 | 38.1 | 180 | 343 | 34.4 |
| Head and Neck | 546 | 4293 | 11.3 | 346 | 3177 | 9.8 |
| Skin | 1468 | 3910 | 27.3 | 1287 | 3427 | 27.3 |

### 2.6 Statistical Analysis Methods

The goal of the algorithms is to produce a well-calibrated prediction of the probability that a patient has cancer. The type of model required is a probabilistic classifier—a model that predicts the probabilities of a given patient belonging to one of several distinct classes.

The development set was used to identify appropriate models and calibration methods and to tune the hyperparameters for those models. Methods and hyperparameters were compared using 5-fold cross-validation. This was concluded and results locked down before validation.

The model structure selected using the development set is a combination of a gradient boosting method, followed by polynomial logistic regression (i.e. a modified version of Platt scaling) to calibrate the resulting predictions. Gradient boosting was chosen for a number of pragmatic and statistical performance reasons, including statistical performance, ability to handle input variables with wildly different distributions (eg tumour markers vs blood counts), an inbuilt method for handling missing data, and modest computational load.

Prior to any analysis variables were selected based on: cost and relevance, availability in NHS pathology labs and prior knowledge from medical literature that they might reasonably be expected to contain some cancer-relevant information. Variable selection in the statistical sense (i.e. using the development data set) was not carried out and the gradient boosting algorithm used in this work is able to down weight any input variables which are of lesser statistical importance (in terms of contribution to making good predictions).

The validation set was used to validate the locked-down algorithms. After this no changes were made to the algorithms, results are presented below.

## 3 Results

Figure 1 shows a CONSORT flow diagram for this work.

**Figure 1:**
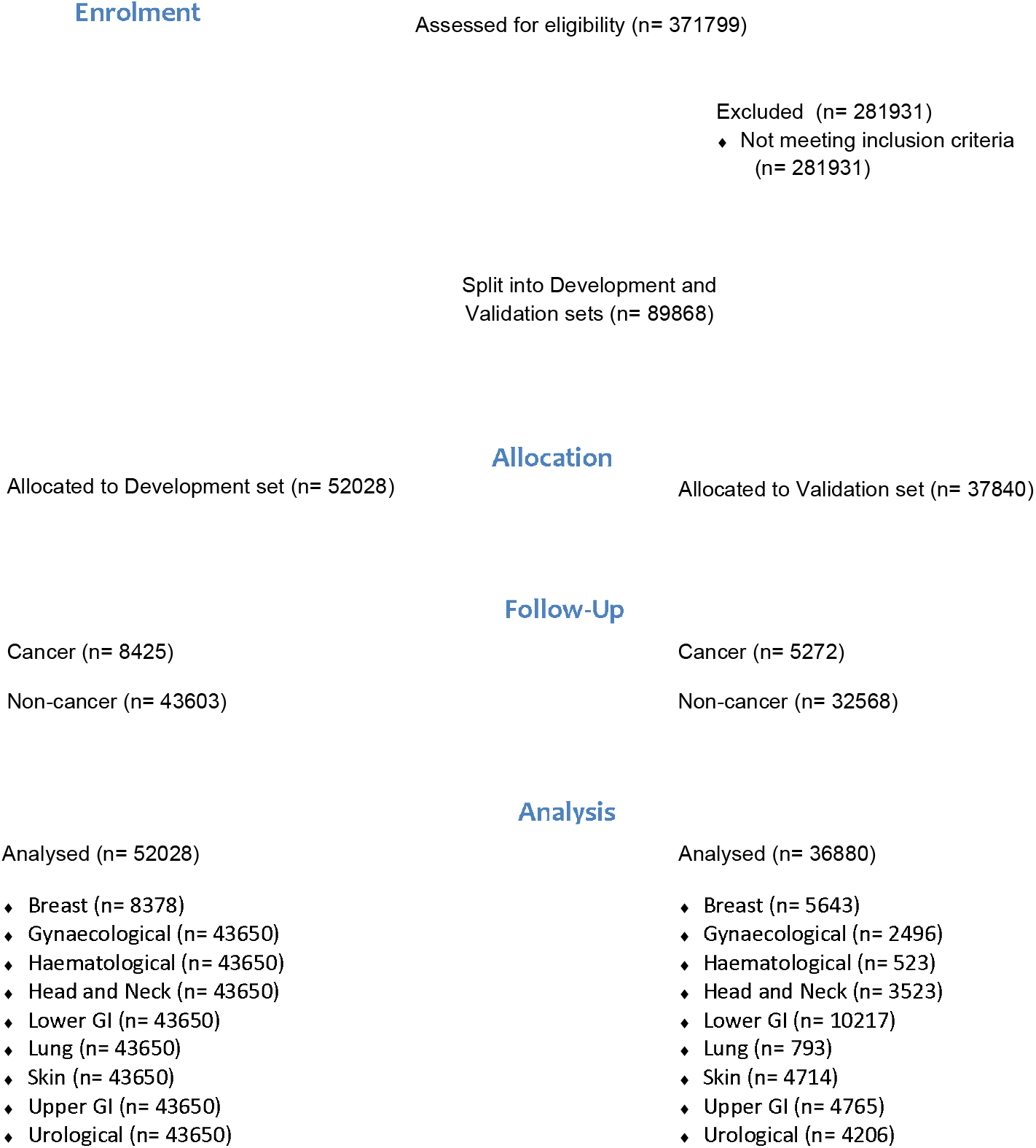
CONSORT flow diagram for this work.

Tables 1 and 2 show the total number of cases per pathway, and the number of those cases meeting the inclusion criteria. Tables 3 and 4 show the age and sex demographics of the included patients, by pathway and by development/validation set.

**Table 3:**
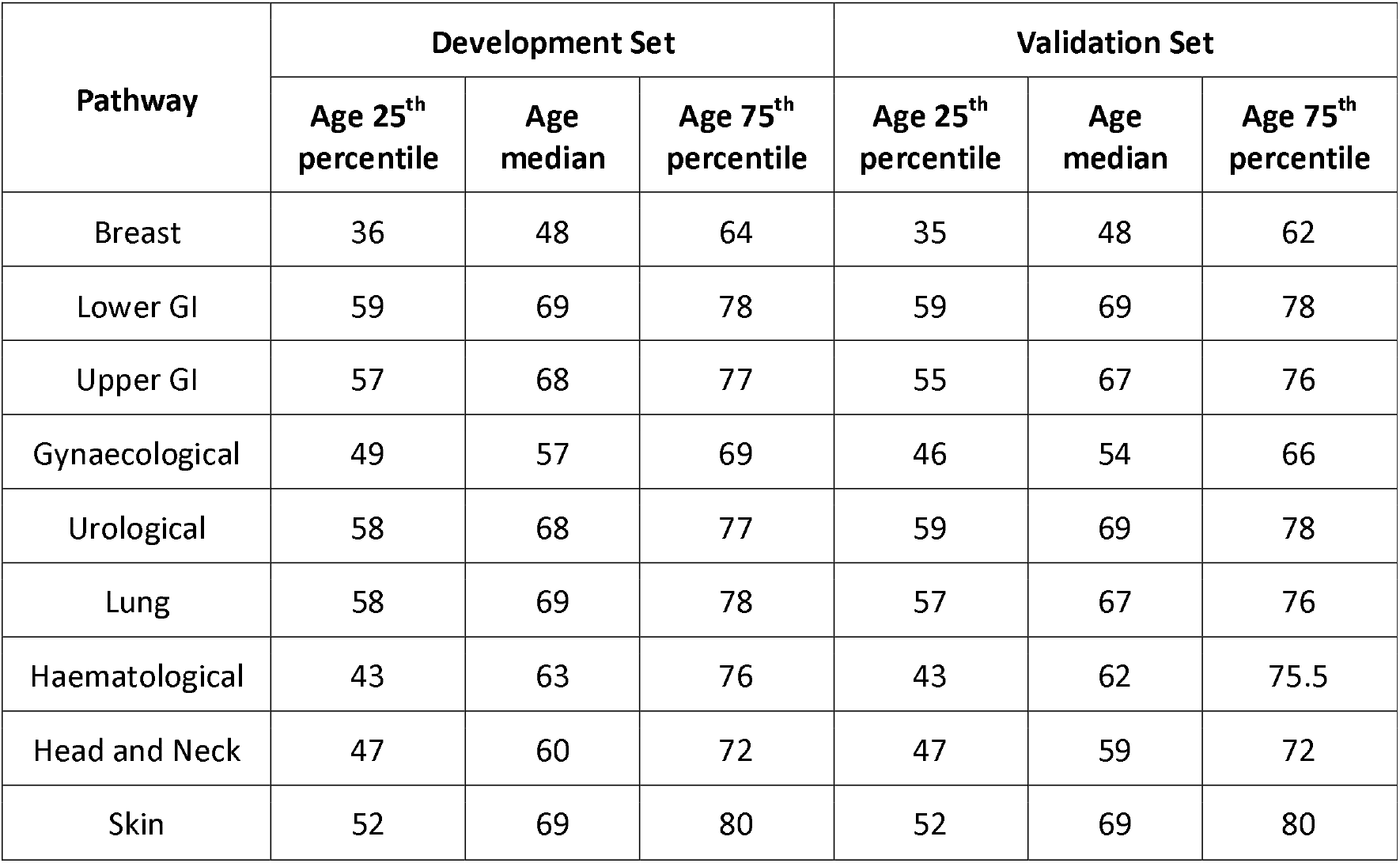
Age Demographics

**Table 4:** Sex Demographics

| **Pathway** | **Development Set** |  | **Validation Set** |  |
| --- | --- | --- | --- | --- |
|  | **# Female (%)** | **# Male (%)** | **# Female (%)** | **# Male (%)** |
| Breast | 7345 (87.67) | 1033 (12.33) | 5146 (91.19) | 497 (8.82) |
| Lower GI | 6889 (54.42) | 5769 (45.58) | 5529 (54.12) | 4688 (45.88) |
| Upper GI | 3346 (55.96) | 2633 (44.04) | 2746 (57.63) | 2019 (42.37) |
| Gynaecological | 3505 (100.00) | 0 (0.00) | 2495 (99.96) | 1 (0.04) |
| Urological | 1700 (26.10) | 4813 (73.90) | 904 (21.49) | 3302 (78.51) |
| Lung | 947 (45.82) | 1120 (54.19) | 363 (45.78) | 430 (54.22) |
| Haematological | 506 (47.87) | 551 (52.13) | 227 (43.40) | 296 (56.60) |
| Head and Neck | 2755 (56.93) | 2084 (43.07) | 2080 (59.04) | 1443 (40.96) |
| Skin | 2924 (54.37) | 2454 (45.63) | 2614 (55.45) | 2100 (44.55) |

Table 5 shows test performance characteristics for nine urgent referral pathways for use-case 1 (rule-out). The goal here is to successfully identify 20% of non-cancer patients (a specificity of 0.2) who are at very low risk of cancer, so that other possible causes of their symptoms can be considered rather than continuing with a 2WW referral.

**Table 5:**
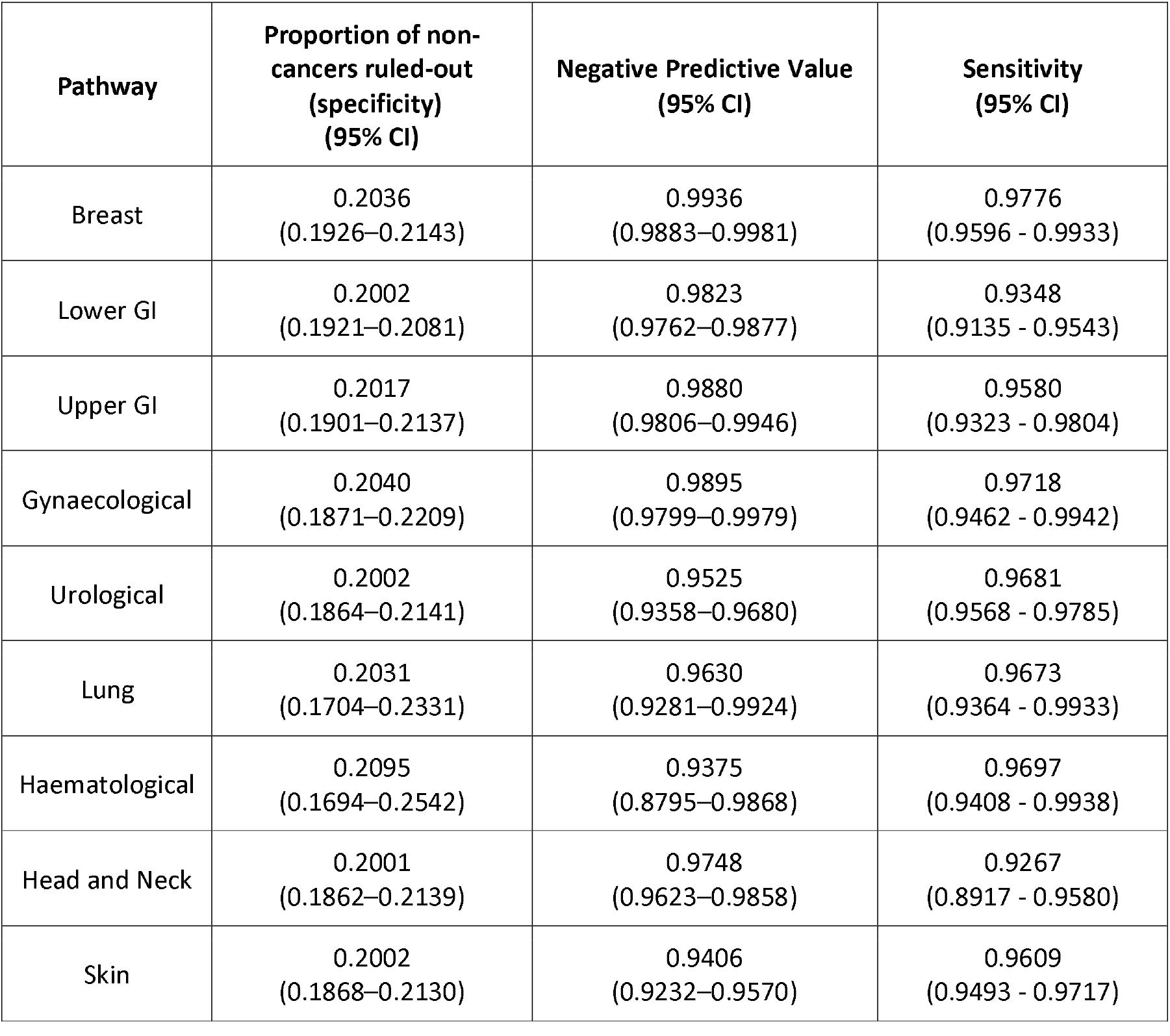
20% Rule-out

Table 6 shows test performance characteristics for use-case 2 (triage), to identify patients at higher risk of cancer who would be considered for priority through the urgent referral pathway. The goal here is to successfully red-flag 90% of cancer cases (a sensitivity of 0.9) for priority investigation.

**Table 6:** 90% Cancer rule-in

| Pathway | Proportion of non-cancers ruled-out<br>(i.e. not red-flagged)<br>(specificity)<br>(95% CI) | Positive Predictive Value<br>(95% CI) |
| --- | --- | --- |
| Breast | 0.4582<br>(0.4450–0.4715) | 0.0890<br>(0.0793 - 0.0991) |
| Lower GI | 0.2723<br>(0.2637–0.2811) | 0.0642<br>(0.0587 - 0.0697) |
| Upper GI | 0.3363<br>(0.3227–0.3503) | 0.0732<br>(0.0644 - 0.0822) |
| Gynaecological | 0.4674<br>(0.4473–0.4879) | 0.1134<br>(0.0972 - 0.1303) |
| Urological | 0.3548<br>(0.3379–0.3710) | 0.3044<br>(0.2878 - 0.3208) |
| Lung | 0.3625<br>(0.3238–0.3987) | 0.2541<br>(0.2178 - 0.2906) |
| Haematological | 0.4330<br>(0.3807–0.4849) | 0.4249 (0.3722 - 0.4759) |
| Head and Neck | 0.2733<br>(0.2579–0.2885) | 0.0804<br>(0.0703 - 0.0911) |
| Skin | 0.3905<br>(0.3745–0.4068) | 0.3230<br>(0.3067 - 0.3392) |

## 4 Discussion

### Summary of main findings

This paper reports the development and validation of a set of statistical machine learning algorithms based on routine laboratory blood measurements that can predict cancer outcomes for symptomatic patients referred urgently from primary care for possible cancer diagnosis.

Each algorithm is trained and validated as a test to provide decision support for one of the nine NHS 2WW pathways. Each test produces a calibrated probability that the patient on that 2WW pathway has any type of cancer. These calibrated probabilities can be used in a range of clinical contexts; in this paper we consider two principal use-cases. In use-case 1, the tests are used to rule-out patients whose risk of cancer is very low, allowing clinicians to identify patients for whom investigations of possible non-cancer causes of their symptoms might be more appropriate. In use-case 2, higher-risk patients are red-flagged so that their onwards journey through the 2WW pathway can be expedited.

Table 5 shows relevant test performance characteristics for use-case 1. With a goal of 20% rule-out and corresponding Negative Predictive Values and Sensitivity, which respectively give the proportion of test-negative results which are correct (i.e. non-cancer cases) and the proportion of cancer cases that are correctly identified as cancer.

Table 6 shows relevant test performance characteristics for use-case 2. Assuming a goal of correctly red-flagging 90% of the cancer cases and presenting the proportion of non-cancer cases that are correctly not red-flagged.

More test performance characteristics can be found in Supplementary Tables S1 and S2.

Figure 2 shows an example of stratification via a test, compared with the existing standard care pathway. In this example, 500 patients present to the breast pathway, which is overloaded and only able to see 400 of these patients within two weeks of their referral. The standard care pathway is modelled as first-come first-served, and so the proportion of patients with cancer is the same in the patients seen and the patients not seen. Using the test for stratification, the patients are stratified into high, medium and low-risk groups. Patients are then seen in risk order - in this example, all of the high-risk patients are seen, and some of the medium-risk patients are seen. Under stratification, far more of the patients with cancer are seen, and of the patients not seen, a far smaller proportion have cancer. An interactive version of this is available at https://www.pinpointdatascience.com/patient-test-stratification

**Figure 2:**
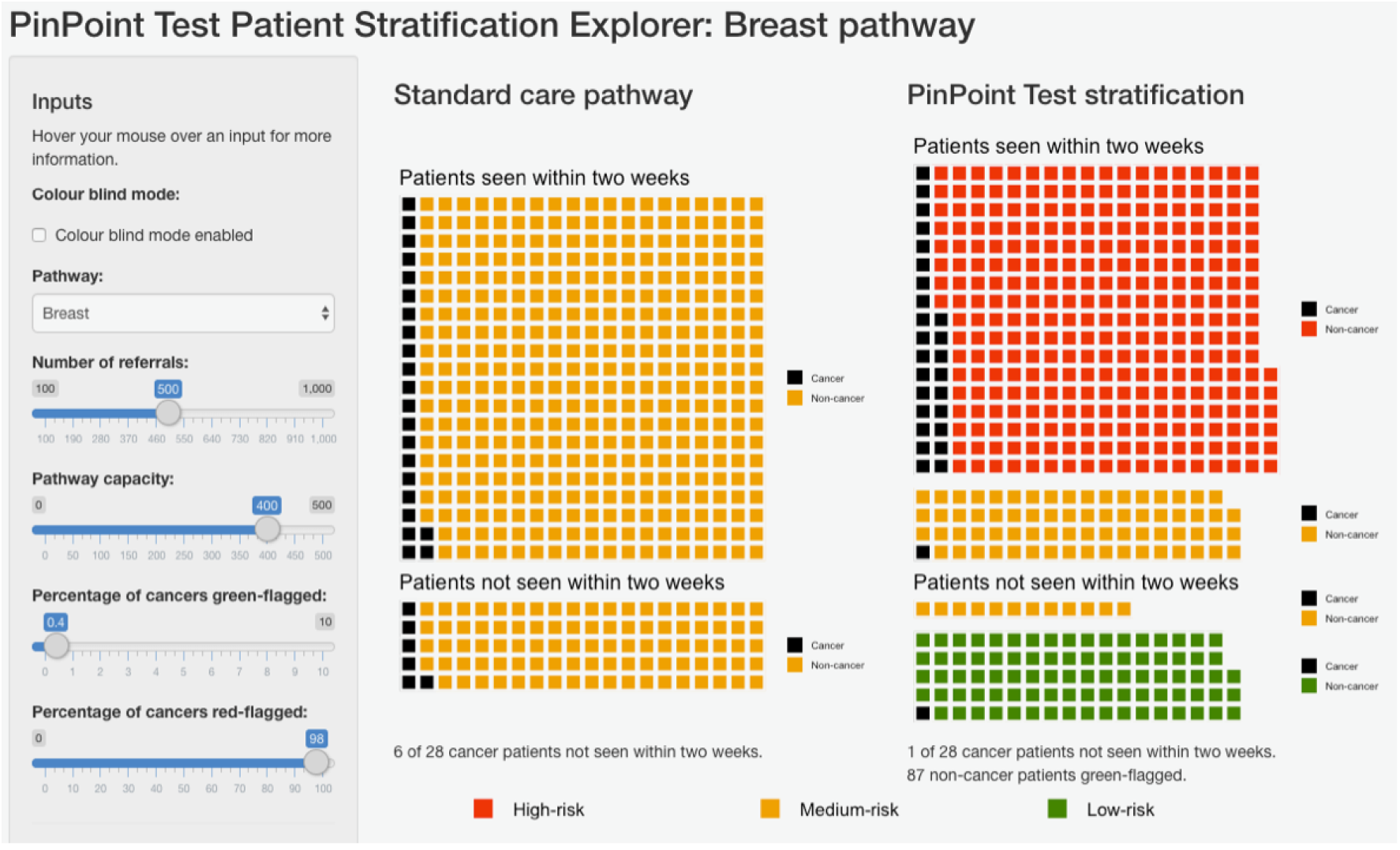
shows stratification of patients on the 2WW breast pathway using the relevant algorithm presented in this work, compared to the standard care pathway. Given an urgent care pathway where the number of referrals exceeds the pathway capacity to see patients within two weeks, use of the test to stratify patients into risk categories (right) leads to a larger proportion of patients with cancer being seen when compared to the standard care pathway (left), in which patients are seen on a first-come, first-served basis. Patients highlighted in red are identified as being at high-risk for cancer (red-flagged), so can be expedited for further diagnostic testing. Patients highlighted in green are identified as being at very low risk for cancer (green-flagged), allowing for initial management in primary care rather than immediate referral to secondary care. The sliders on the left-hand side show the number of referrals, the number of patients that the pathway can handle in a given time-frame (the pathway capacity), the percentage of cancers which are green-flagged (i.e. setting a very low false negative rate), and the percentage of cancers that are red-flagged (i.e. identifying cases with high-risk, so that they can be expedited for further diagnostic testing). Collectively, this represents a possible approach to using the algorithms to improve the triage of patients referred to a 2WW pathway. An interactive version of this is available at https://www.pinpointdatascience.com/patient-test-stratification

### 4.1 Discussion of main findings within the context of the literature

This work is novel, innovative, and potentially of huge importance for the management of patients referred urgently for suspected cancer. The tests are based upon a panel of routine blood measurements that: are already in common usage in NHS laboratories; work across a range of cancers; can easily be integrated with existing NHS systems. The tests have already been integrated with Mid-Yorkshire Hospitals NHS Trust Laboratory systems.

The tests can both identify patients at higher risk of cancer, such that they can be prioritised for assessment and diagnostic investigations, while also identifying a significant proportion of patients at very low risk who may not need further investigation for suspected cancer. Patients in both groups stand to benefit, either from expedited testing, or from not being exposed to iatrogenic harm and unnecessary cancer worries. The tests can be set at different thresholds in different cancers and within different health settings, making them responsive to local needs, capacity and priorities. COVID has reduced diagnostic capacity and efficiency, this test could be an effective and rapid solution at this time of crisis.

An important practical note is that the criteria for 2WW changed in 2015, reducing the risk threshold warranting an urgent referral from 5% PPV to 3% PPV (i.e. towards the end of the development cohort timeframe). The validation results therefore encompass this change in clinical practice, suggesting a certain robustness to those results.

#### Strengths

The principal strengths of this work are:

⍰ It is based on well-validated, low-cost clinical assays already available at scale in NHS pathology laboratories.
⍰ The tests could therefore be deployed across the UK very rapidly, with no additional hardware requirements.
⍰ The tests are CE marked and are currently undergoing service evaluation in the West Yorkshire and Harrogate Cancer Alliance.
⍰ The performance estimates are conservative due to missing data and the historical nature of the blood measurements; prospective evaluation will not suffer from these drawbacks
⍰ Even biomarkers with limited individual performance are of value in this approach if they contribute complementary information
⍰ The algorithms are designed to be flexible, allowing thresholds to be changed according to clinical need, for example Use-Case 2 during the COVID-19 pandemic
⍰ The large numbers reported, the robust analysis and reporting in line with TRIPOD and PROBAST.^11,12^
⍰ There is the potential to improve performance using the pipeline of new biomarkers being developed for diagnostic, predictive or prognostic purposes.

#### Limitations

The principal limitations of this work are:

⍰ That the development and validation was done only in one centre.
⍰ There is a possible source of bias, in that the subset of patients who had retrospective blood data may not be representative of the overall 2WW cohort.
⍰ We have only reported the validation on a retrospective sample; a prospective evaluation is needed.
⍰ The validation set meets the defined sample size criteria (1500 total cases) for 7 of the 9 2WW. 95% CI are provided for all results to make clear the level of uncertainty present due to sample sizes.
⍰ The remaining (smaller) 2WW pathways as recorded in the clinical data were also considered (Testicular, Brain/CNS, Sarcomas, Children’s Cancer, Acute Leukaemia, HPB, Thyroid Cancer, Renal, other cancer), but we did not develop algorithms for these as the available sample sizes were judged too small to train and validate effective models.

### 4.2 Implications for policy research and practice

Until we have undertaken a prospective evaluation of the performance of the algorithms it is not possible to predict how this will be used. However, we do envisage use of the tool, as part of clinical triage, to both prioritise those at higher levels of risk and de-prioritise those at the very lowest levels of risk, in conjunction with appropriate safety netting. We also need to fully understand the views of patients, clinicians, and commissioners on the acceptability and utility of the tests.

## Data Availability

The data will not be made available to others, as it is de-identified NHS patient data.

## Acknowledgements

We thank the following people and organisations for their contributions:

⍰ Emmylou Laird and Claire Eckert for their help with numerous aspects of this work
⍰ The Leeds Institute for Data Analytics, in particular Phil Chambers, for all their support in providing secure computing facilities.
⍰ The pathology and health informatics staff of Leeds Teaching Hospitals Trust and Mid-Yorkshire Hospitals NHS Trust for their help.
⍰ The X-Lab team for help with deployment into the NHS
⍰ The board members and partner organisations of the Leeds Centre for Personalised Medicine and Health and the Leeds Academic Health Partnership.
⍰ The NIHR Leeds MIC and CRUK CANTEST PPI groups, especially Pete Whetstone and Margaret Johnson.
⍰ We thank HealthWatch Leeds and Healthwatch Kirklees for patient consultation forums

## Authors’ contributions

RS, MM, RN, GH, RF and SD conceptualised the study, and led on the initial protocol development. GT, RF, NPS, BS and PS contributed towards funding applications and protocol refinement. RS, MN, KL and JS developed the software and algorithms, performed the data analysis and completed the CE marking process, with clinical input from RN, SD, NS, GH and PS and methodological input from BS, CJ and MM. GH led on the provision of de-identified data, assisted by CJ and RF. RF oversaw project management. All authors contributed to the interpretation of the results, writing of the manuscript and approved the final version.

## Ethics statement

Data for the analysis are retrospective and fully de-identified before being released to the study team. The work was carried out under service evaluation with the formal approval of the Leeds Teaching Hospitals Trust R&I and Data Governance Committee, and with the specific approval of the Trust Caldicott Guardian.

## Data availability

The data will not be made available to others, as it is de-identified NHS patient data.

## TRIPOD

This work is reported in accordance with the TRIPOD statement.

