## Supplementary Materials for "Development and validation of multivariable machine learning algorithms to predict risk of cancer in symptomatic patients referred urgently from primary care"

### Test Performance Characteristics

Table S1: Test validation set performance characteristics. Aim: 20% rule-out

| Pathway | Threshold | AUC (95% CI) | NPV (95% CI) | TNR (95% CI) | FNR (95% CI) | Sensitivity (95% CI) | Specificity (95% CI) | PPV (95% CI) |
|---|---|---|---|---|---|---|---|---|
| Breast | 0.0174 | 0.8007 (0.7750 – 0.8255) | 0.9936 (0.9883 – 0.9981) | 0.2036 (0.1926 – 0.2143) | 0.0224 (0.0067 – 0.0404) | 0.9776 (0.9596 – 0.9933) | 0.2036 (0.1926 – 0.2143) | 0.0672 (0.0601 – 0.0747) |
| Lower GI | 0.0343 | 0.6798 (0.6566 – 0.7029) | 0.9823 (0.9762 – 0.9877) | 0.2002 (0.1921 – 0.2081) | 0.0652 (0.0457 – 0.0865) | 0.9348 (0.9135 – 0.9543) | 0.2002 (0.1921 – 0.2081) | 0.0609 (0.0559 – 0.0660) |
| Upper GI | 0.0284 | 0.7323 (0.7008 – 0.7627) | 0.9880 (0.9806 – 0.9946) | 0.2017 (0.1901 – 0.2137) | 0.0420 (0.0196 – 0.0677) | 0.9580 (0.9323 – 0.9804) | 0.2017 (0.1901 – 0.2137) | 0.0653 (0.0576 – 0.0732) |
| Gynaecological | 0.0392 | 0.8124 (0.7779 – 0.8459) | 0.9895 (0.9799 – 0.9979) | 0.2040 (0.1871 – 0.2209) | 0.0282 (0.0058 – 0.0538) | 0.9718 (0.9462 – 0.9942) | 0.2040 (0.1871 – 0.2209) | 0.0852 (0.0732 – 0.0980) |
| Urological | 0.1062 | 0.7590 (0.7414 – 0.7757) | 0.9525 (0.9358 – 0.9680) | 0.2002 (0.1864 – 0.2141) | 0.0319 (0.0215 – 0.0432) | 0.9681 (0.9568 – 0.9785) | 0.2002 (0.1864 – 0.2141) | 0.2751 (0.2609 – 0.2900) |
| Lung | 0.0876 | 0.7376 (0.6938 – 0.7797) | 0.9630 (0.9281 – 0.9924) | 0.2031 (0.1704 – 0.2331) | 0.0327 (0.0067 – 0.0636) | 0.9673 (0.9364 – 0.9933) | 0.2031 (0.1704 – 0.2331) | 0.2249 (0.1934 – 0.2571) |
| Haematological | 0.111 | 0.7589 (0.7152 – 0.8006) | 0.9375 (0.8795 – 0.9868) | 0.2095 (0.1694 – 0.2542) | 0.0303 (0.0062 – 0.0592) | 0.9697 (0.9408 – 0.9938) | 0.2095 (0.1694 – 0.2542) | 0.3612 (0.3166 – 0.4068) |
| Head and Neck | 0.0423 | 0.6996 (0.6649 – 0.7334) | 0.9748 (0.9623 – 0.9858) | 0.2001 (0.1862 – 0.2139) | 0.0733 (0.0420 – 0.1083) | 0.9267 (0.8917 – 0.9580) | 0.2001 (0.1862 – 0.2139) | 0.0755 (0.0657 – 0.0852) |
| Skin | 0.0851 | 0.7220 (0.7057 – 0.7378) | 0.9406 (0.9232 – 0.9570) | 0.2002 (0.1868 – 0.2130) | 0.0391 (0.0283 – 0.0507) | 0.9609 (0.9493 – 0.9717) | 0.2002 (0.1868 – 0.2130) | 0.2796 (0.2656 – 0.2939) |

Table S2: Test validation set performance characteristics. Aim: 90% rule-in

| Pathway | Threshold | AUC (95% CI) | NPV (95% CI) | TNR (95% CI) | FNR (95% CI) | Sensitivity (95% CI) | Specificity (95% CI) | PPV (95% CI) |
|---|---|---|---|---|---|---|---|---|
| Breast | 0.029 | 0.8007 (0.7746 – 0.8256) | 0.9875 (0.9830 – 0.9916) | 0.4582 (0.4450 – 0.4715) | 0.0990 (0.0678 – 0.1337) | 0.9010 (0.8663 – 0.9322) | 0.4582 (0.4450 – 0.4715) | 0.0890 (0.0793 – 0.0991) |
| Lower GI | 0.041 | 0.6798 (0.6565 – 0.7029) | 0.9799 (0.9745 – 0.9850) | 0.2723 (0.2637 – 0.2811) | 0.1006 (0.0754 – 0.1262) | 0.8994 (0.8738 – 0.9246) | 0.2723 (0.2637 – 0.2811) | 0.0642 (0.0587 – 0.0697) |
| Upper GI | 0.041 | 0.7323 (0.7012 – 0.7625) | 0.9831 (0.9763 – 0.9893) | 0.3363 (0.3227 – 0.3503) | 0.0992 (0.0641 – 0.1389) | 0.9008 (0.8611 – 0.9359) | 0.3363 (0.3227 – 0.3503) | 0.0732 (0.0644 – 0.0822) |
| Gynaecological | 0.05 | 0.8124 (0.7768 – 0.8462) | 0.9828 (0.9746 – 0.9900) | 0.4674 (0.4473 – 0.4879) | 0.1073 (0.0640 – 0.1553) | 0.8927 (0.8447 – 0.9360) | 0.4674 (0.4473 – 0.4879) | 0.1134 (0.0972 – 0.1303) |
| Urological | 0.148 | 0.7590 (0.7417 – 0.7762) | 0.9191 (0.9035 – 0.9336) | 0.3548 (0.3379 – 0.3710) | 0.0996 (0.0818 – 0.1183) | 0.9004 (0.8817 – 0.9182) | 0.3548 (0.3379 – 0.3710) | 0.3044 (0.2878 – 0.3208) |
| Lung | 0.134 | 0.7376 (0.6939 – 0.7796) | 0.9431 (0.9120 – 0.9702) | 0.3625 (0.3238 – 0.3987) | 0.0915 (0.0482 – 0.1392) | 0.9085 (0.8608 – 0.9518) | 0.3625 (0.3238 – 0.3987) | 0.2541 (0.2178 – 0.2906) |
| Haematological | 0.189 | 0.7589 (0.7143 – 0.7999) | 0.9118 (0.8633 – 0.9509) | 0.4330 (0.3807 – 0.4849) | 0.0909 (0.0506 – 0.1412) | 0.9091 (0.8588 – 0.9494) | 0.4330 (0.3807 – 0.4849) | 0.4249 (0.3722 – 0.4759) |
| Head and Neck | 0.047 | 0.6996 (0.6648 – 0.7339) | 0.9751 (0.9644 – 0.9847) | 0.2733 (0.2579 – 0.2885) | 0.0991 (0.0619 – 0.1393) | 0.9009 (0.8607 – 0.9381) | 0.2733 (0.2579 – 0.2885) | 0.0804 (0.0703 – 0.0911) |
| Skin | 0.141 | 0.7220 (0.7060 – 0.7380) | 0.9236 (0.9100 – 0.9367) | 0.3905 (0.3745 – 0.4068) | 0.0999 (0.0829 – 0.1175) | 0.9001 (0.8825 – 0.9171) | 0.3905 (0.3745 – 0.4068) | 0.3230 (0.3067 – 0.3392) |

**Clinical Utility Plots**

Figure S1 shows negative predictive value (NPV) against the specificity, i.e. the proportion of patients ruled out, for each pathway. Bootstrap resampling with replacement with 1000 bootstraps was used to generate 95% and 68% confidence intervals on NPV. The solid line marks the median, the dark grey band indicates the 68% confidence interval, and the light grey band indicates the 95% confidence interval.

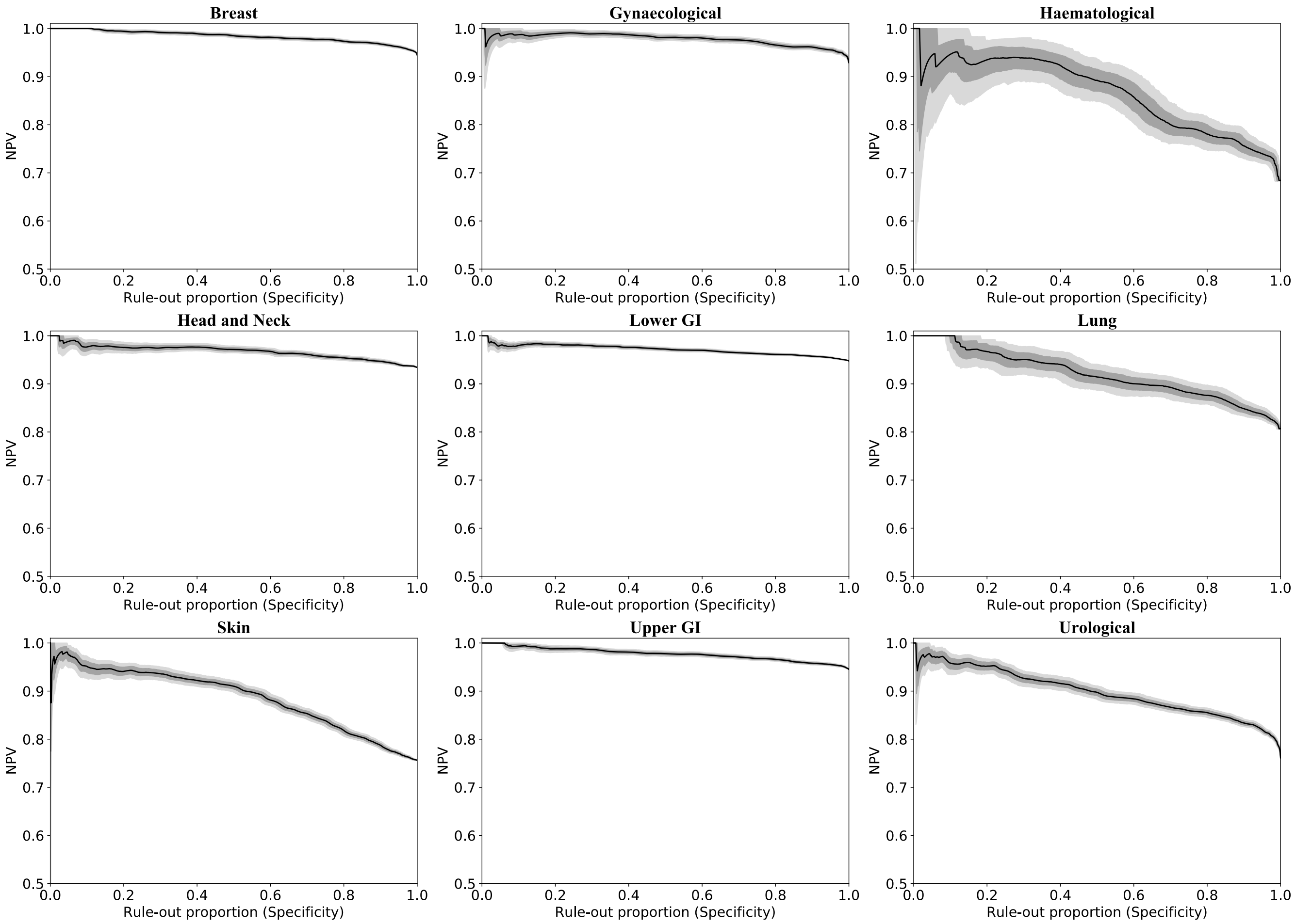

Figure S1: Plots of Negative Predictive Ability against specificity for each pathway. Light and dark grey bands indicate 68% and 95% confidence intervals. See text for details.

### Calibration

Figure S2 shows calibration curves for validation set predictions by the algorithms for each pathway, calculated using equal occupancy bins. The error bars show the 95% binomial proportion confidence interval, calculated using the Wilson score with continuity correction. The log loss for each pathway is also included.

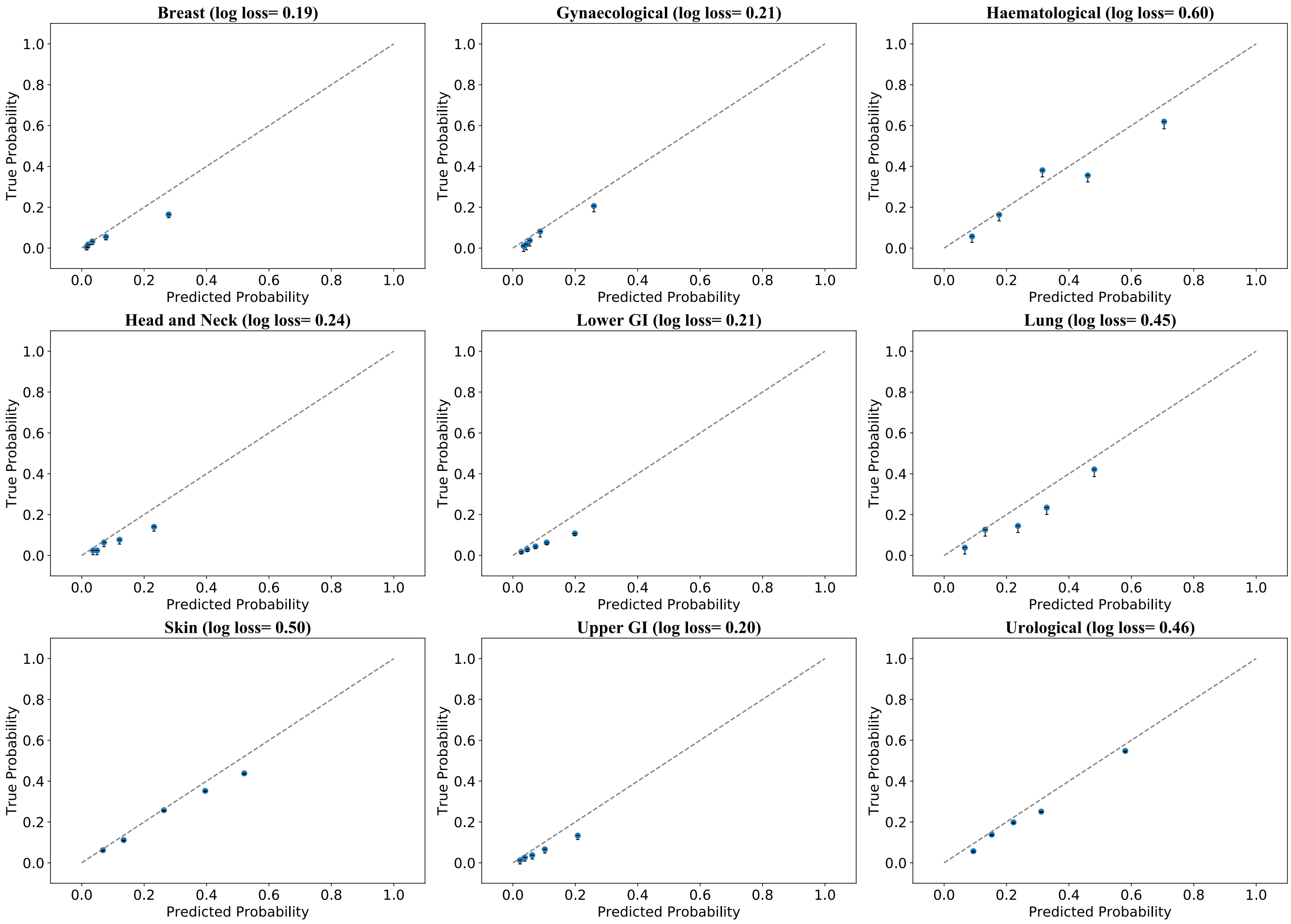

Figure S2: Plots of calibration curves per pathway. Dashed grey line indicates perfect calibration. See text for details.

**Univariate Analyses**
Validation set predicted probabilities were generated using the nine algorithms. For each input data feature, ROC AUCs were calculated for cases restricted to those for which the feature data was available, whereby the feature was used as the predictor and the binary cancer flag as the outcome. ROC AUCs were also calculated using the probabilities predicted by the algorithm, with identical restriction of cases applied to allow direct comparison. The difference between the algorithm ROC AUC and the single-feature ROC AUC was then calculated for each feature, ΔAUC.

Using this process, ΔAUCs were calculated for each feature and each pathway-specific algorithm. Bootstrap resampling with replacement with 10000 bootstraps was used to generate 95% confidence intervals on ΔAUC, where both the algorithm ROC AUC and single-feature ROC AUC were calculated on the same bootstrap samples.

Figure S3 shows the median ΔAUCs as black circles with 95% confidence intervals, for each feature and each pathway. Any features with data for less than one hundred patients for a given pathway were removed from the plot for that pathway. Arrows indicate that a confidence interval extends outside the plot area, in the direction of the arrow. The number of cancers and the number of cases were annotated for each feature at the bottom of the plot area. These are in the format "# cancers/# cases". An asterisk was appended to feature names for which the 95% confidence interval does not intersect the line $\Delta AUC = 0$. The feature names are assigned according to the category into which the blood test falls—"FBC" for blood counts, "Bio" for biochemistry, and "TM" for tumour markers—with numbers assigned arbitrarily but consistently across the subplots.

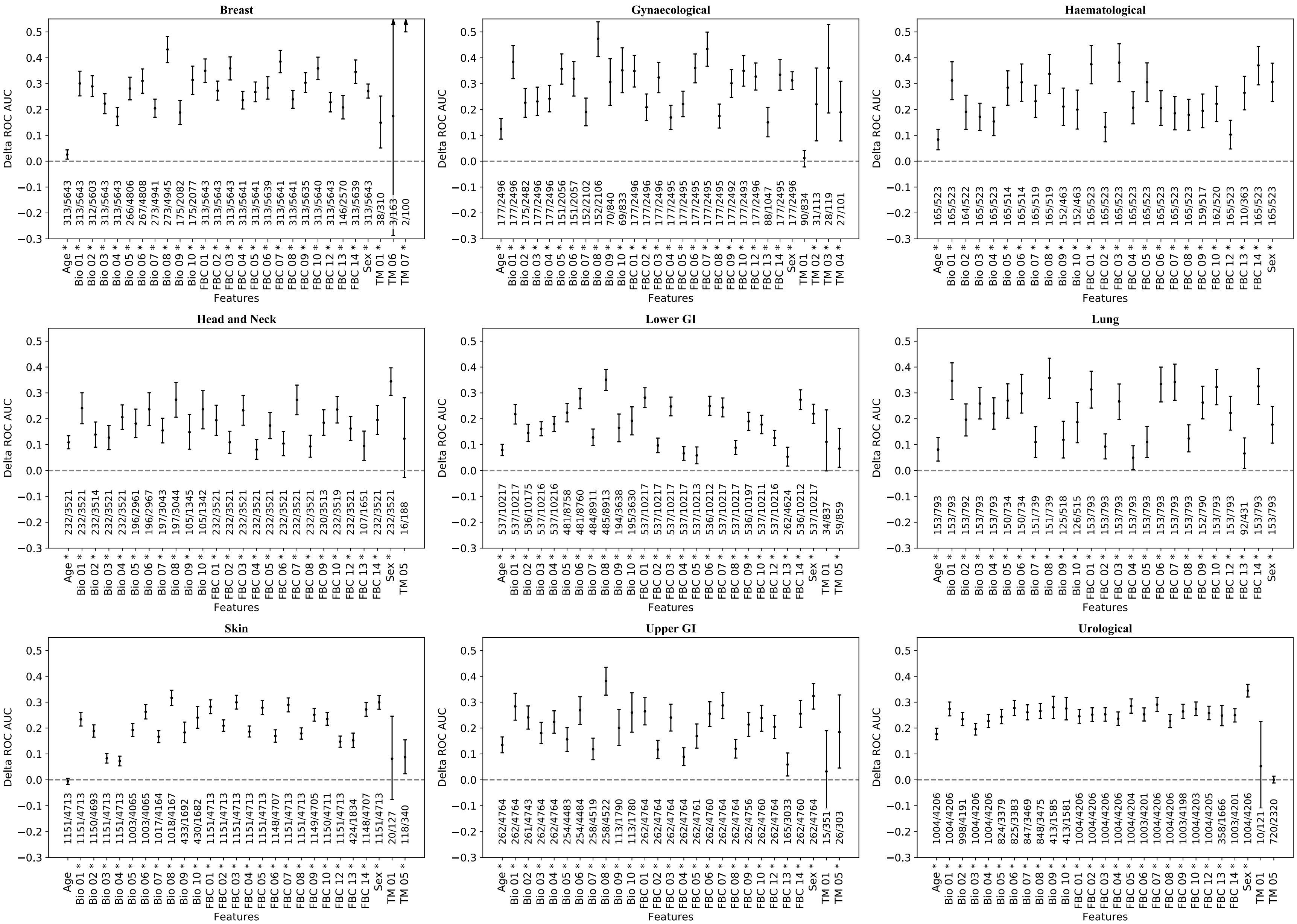

Figure S3: Plots of ΔAUC per feature per pathway. See text for details.

**ICD-10 Codes**

Table S3: ICD-10 codes designated as “cancer” for the algorithms

| ICD-10 code | ICD-10 text |
|---|---|
| C00-C14 | Malignant neoplasms of lip, oral cavity and pharynx |
| C15-C26 | Malignant neoplasms of digestive organs |
| C30-C39 | Malignant neoplasms of respiratory and intrathoracic organs |
| C40-C41 | Malignant neoplasms of bone and articular cartilage |
| C43-C44 | Melanoma and other malignant neoplasms of skin |
| C45-C49 | Malignant neoplasms of mesothelial and soft tissue |
| C50-C50 | Malignant neoplasm of breast |
| C51-C58 | Malignant neoplasms of female genital organs |
| C60-C63 | Malignant neoplasms of male genital organs |
| C64-C68 | Malignant neoplasms of urinary tract |
| C69-C72 | Malignant neoplasms of eye, brain and other parts of central nervous system |
| C73-C75 | Malignant neoplasms of thyroid and other endocrine glands |
| D00 | Carcinoma in situ of oral cavity, oesophagus and stomach |
| D01 | Carcinoma in situ of other and unspecified digestive organs |
| D02 | Carcinoma in situ of middle ear and respiratory system |
| D03 | Melanoma in situ |
| D04 | Carcinoma in situ of skin |
| D05 | Carcinoma in situ of breast |
| D07 | Carcinoma in situ of other and unspecified genital organs |
| D09 | Carcinoma in situ of other and unspecified sites |

Table S4: ICD-10 codes designated as “benign” for the algorithms

| ICD-10 code | ICD-10 text |
|---|---|
| D06 | Carcinoma in situ of cervix uteri |
| D10-D36 | Benign neoplasms |
| D37-D48 | Neoplasms of uncertain or unknown behaviour |
